# Determining the role of lifestyle factors in healthy cognitive preservation in SuperAgers: A systematic review

**DOI:** 10.1101/2024.11.15.24317375

**Authors:** Philippa Watson, Megan Smith, Ivan Koychev, John Gallacher, Sarah Bauermeister

**Affiliations:** Department of Psychiatry, University of Oxford

## Abstract

**Background:** A subset of older adults, known as SuperAgers (SAs), exhibit exceptional resilience to these effects, displaying cognitive abilities at the same level or exceeding those 20-30 years younger. To date however, there is little understanding as to which factors may be responsible for SA enhanced cognitive abilities in old age. This systematic review aimed to identify and evaluate the evidence for an association between lifestyle factors and SA status.

**Methods:** A systematic literature search in accordance with the PRISMA guidelines was conducted across MedLine, Embase, Ovid, Global Health, APA PsychArticles, and PsycINFO from inception to 06/11/23 of studies investigating the link between SA status and one or more lifestyle factors.

**Results:** A systematic search identified 13 studies which met the inclusion criteria. Eleven investigated the relationship between two or more lifestyle factors with the most common lifestyle factor examined being mental health followed by physical activity, social engagement and smoking. Mixed results were observed across the included studies while social engagement and mental health emerged as the most likely lifestyle factors to be positively associated with SA status, there was considerable heterogeneity in the measures used to assess different lifestyle factors.

**Conclusion:** No clear conclusions could be drawn as to which lifestyle factors are associated with SA status due to scarcity of studies and heterogeneity in the measurement of different lifestyle factors.

## 1. Introduction

Cognitive ageing is highly heterogeneous and synonymous with the ageing process. While some experience cognitive impairment, commonly conceptualised through diagnoses such as mild cognitive impairment (MCI) or various dementias, including Alzheimer’s disease (AD), there exists a group of adults who exhibit superior cognitive abilities well into older age (Harrison et al., 2012; Rogalski et al., 2013; Yu et al., 2020). Whilst cognitive impairment in older adults has been studied extensively over the past few decades, far fewer studies have examined the opposite side of the spectrum: older adults with maintained or superior cognitive abilities (Petersen et al., 2014; Yu et al., 2020). There have been multiple efforts in the literature to categorise this group, with the most used term being ‘SuperAgers’ (Harrison et al., 2012; Rogalski et al., 2013). Hence, a SuperAger (SA) is defined as a person who appears to resist age-related cognitive decline and instead exhibits, for example, superior memory performance by scoring as high or higher as healthy adults 20–30 years younger (Garo-Pascual et al., 2023; Harrison et al., 2012; Maccora et al., 2021).

Existing research on SAs, including a recent systematic review on brain resilience in SAs (de Godoy et al., 2021), has identified specific brain regions associated with cortical preservation. Specifically, studies have shown that SAs have greater cortical thickness in brain areas such as the hippocampus (Dekhtyar et al., 2017; Harrison et al., 2018; Sun et al., 2016; Yang et al., 2016), anterior cingulate cortex (Gefen et al., 2015; Harrison et al., 2012; Sun et al., 2016), medial prefrontal cortex (Harrison et al., 2018; Sun et al., 2016; Yang et al., 2016), comprehending structures of the default mode network (relating to episodic memory function), and insular (part of the salience network associated with attention and executive processes in encoding and retrieval) (de Godoy et al., 2021; Harrison et al., 2018; Sun et al., 2016; Yang et al., 2016). Research has also shown that SAs are less likely to carry the APOE-E4 allele and have a lower frequency of AD pathology (Rogalski et al., 2013). Overall, this research indicates that SAs may possess distinctive brain physiology, which is noteworthy given that age-related cortical atrophy typically begins as early as young adulthood and is well-documented across the lifespan (Cook Maher et al., 2022; Fotenos et al., 2005; Resnick et al., 2003).

However, a comprehensive understanding of modifiable factors contributing to cognitive preservation in SuperAgers is still lacking (Garo-Pascual et al., 2023; Maccora et al., 2021). Identifying any modifiable factors that may enable SuperAgers to maintain their healthy cognitive abilities is of notable importance, as this may provide important insights about mechanisms of risk and resilience. It can be theorised however, that the same factors that may be important in delaying the progression to MCI and AD might also play a role in maintaining cognitive health in SAs. Such insights could inform the development of preventative treatments for cognitive decline and dementia, potentially alleviating its public health burden (Anstey & Christensen, 2000; Burke et al., 2019).

Cumulative epidemiological evidence strongly suggests that a healthy lifestyle is related to cognitive health in later life. Regular exercise (Erickson et al., 2022; Gill et al., 2015; Horder et al., 2018; Omura et al., 2020), moderate alcohol consumption (Jeon et al., 2023; Zarezadeh et al., 2024), smoking cessation (Amini et al., 2021; Linli et al., 2022), high social activity engagement (Kelly et al., 2017; Liu et al., 2019; Sala et al., 2019), among other lifestyle factors have all been shown across a multitude of studies to lower the rate of memory decline in older adults and reduce the risk of dementia. For instance, in a recent comprehensive report on dementia prevention, intervention, and care, Livingston and colleagues (2024) reported that addressing 14 risk factors, including smoking, physical inactivity, and social inactivity, could delay up to 40% of all dementia diagnoses (Livingston et al., 2024). To summarise, this existing body of evidence suggests that lifestyle factors may play an important role in cognition preservation, making it a worthwhile research area to investigate when considering what may be associated with SA’s maintained or enhanced cognitive abilities.

The purpose of this review was to conduct a systematic investigation of existing SA literature. The objective is to collate key findings on the role of lifestyle factors which may be important to preserve healthy cognitive abilities, highlighting avenues for future research and intervention. Each lifestyle factor and its relationship to SA status is considered in turn, with a focus given to common measures of lifestyle engagement (e.g., physical activity, smoking, social engagement, etc.) previously identified as modifiable risk factors for cognitive change (Livingston et al., 2024).

## 2 Methods

### 2.1 Eligibility criteria

A systematic search of the literature, including studies in English and no limit on publication date, was made following the 2010 Preferred Reporting Items for Systematic Reviews and Meta-Analyses (PRISMA) recommendations. The following inclusion criteria were applied: articles published in English; articles that evaluated engagement in one or more lifestyle factors (e.g., diet, physical activity, smoking, social engagement); articles that examined individuals aged 65 and over in groups with at least one group classified as SA or another terminology with a similar concept (older adults with exceptional cognitive performance). The exclusion criteria were: studies not utilising original data, conference abstracts, book chapters, theses, and review papers.

### 2.2 Search strategy

The databases MedLine, Embase, Ovid, Global Health, APA PsychArticles, and PsycINFO were searched on 06/11/23, with no time restriction to identify relevant articles using search terms relating to SAs: (SuperAge*” OR “Resilient ager*” OR “successful memory ageing*” OR “successful memory ageing*” OR “Super-cognition*” OR “Superior cognition*” OR “Exceptional memory*” OR “Superior memory performance*” OR “successful cognitive ageing*” OR “successful cognitive ageing*”). As for PsycINFO, the following filters were employed: “publication type: peer-reviewed”; “population group: human”; “age group: 65 yrs & older.” The retrieval of the studies and the removal of duplicates were performed using the Rayyan tool (Ouzzani et al., 2016).

### 2.3 Study Selection

Initially, titles and abstracts were screened against the criteria by one reviewer. Subsequently, two independent reviewers conducted a full-text review. Any disagreements were resolved through consensus with a third reviewer with expertise in neuropsychology and epidemiology.

### 2.4 Data Extraction

For each study, one reviewer extracted the following information into a standardized format: study details (author, publication year, country, design, sample approach); participant information (sample size, average age, gender distribution); statistical methods used, and covariates considered; and lifestyle measures, SA assessment method, and key findings.

### 2.5 Study risk of bias assessment

The risk of bias was assessed using the Quality in Prognosis Studies (QUIPS, Hayden et al., 2013) tool for each included study by the two independent reviewers, with discrepancies resolved through consensus with the third independent reviewer.

### 2.6 Data Synthesis

A narrative synthesis was conducted to summarise the findings of all the studies included in the systematic review. The main reporting on participant characteristics and definitions of SAs are presented in a separate table. The main lifestyle factor findings are sorted and grouped according to the lifestyle measure examined and presented in separate tables.

## 3 Results

### 3.1 Study selection and characteristics

Among the 1697 articles from the primary search 13 met the inclusion criteria (see Figure 1.1 for flow chart of selection process). These 13 studies were conducted across 9 different countries and four different continents; four studies were conducted in the USA with one of these studies examining only African American participants and another only Latinx participants (see Table 1.2 for further particulars). Across the included studies, the following lifestyle factors were investigated: physical activity, social activity engagement, diet, alcohol, smoking, mental health, and sleep (see Table 1.1 for study lifestyle factor inclusion).

**Figure 1.1.**
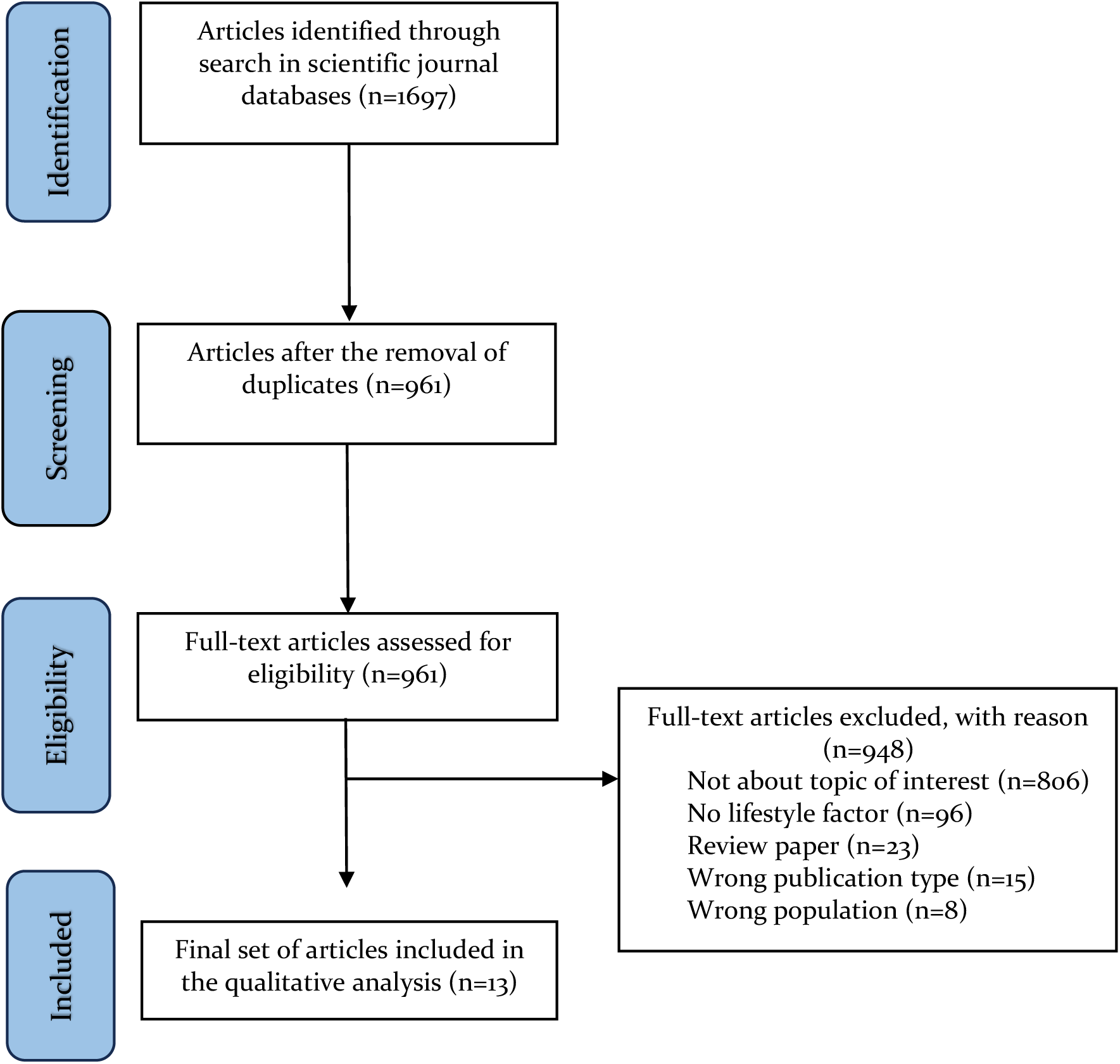
PRISMA flow-chart of search for and selection of included studies.

**Table 1.1.** Lifestyle factors targeted in the included studies.

| Study | Physical Activity | Social Engagement | Diet | Alcohol | Smoking | Mental Health | Sleep |
| --- | --- | --- | --- | --- | --- | --- | --- |
| Calandri and colleagues (2020) | √ | √ | - | - | √ | - | - |
| Cook Maher and colleagues (2022) | - | - | - | - | - | √ | - |
| Dang and colleagues (2019) | - | - | - | - | - | √ | - |
| Doyle and colleagues (2024) | - | - | - | - | √ | √ | - |
| García-Chanes and colleagues (2022) | - | √ | - | - | - | √ | - |
| Garó-Pascual and colleagues (2023) | √ | √ | √ | √ | √ | √ | √ |
| Hermansen and colleagues (2024) | - | - | - | √ | √ | √ | - |
| Maccora and colleagues (2020) | √ | √ | - | √ | √ | √ | √ |
| Saint Martin and colleagues (2017) | √ | √ | - | √ | √ | √ | - |
| Trammell and colleagues (2024) | - | - | - | √ | - | √ | √ |
| Wagner and Grodstein (2022) | √ | - | √ | - | - | - | √ |
| Yang and colleagues (2022) | √ | √ | - | - | - | √ | - |
| Yu and colleagues (2020) | √ | √ | - | √ | √ | √ | √ |
| Total number of lifestyle factors | 7 | 7 | 2 | 6 | 7 | 11 | 5 |

**Table 1.2.** Participant characteristics for included studies.

| Authors, year | Country | Sample Size (No. SAs, No. controls) | % female Total (% female SA) | Study Design (cohort if applicable) | SuperAger Assessment |
| --- | --- | --- | --- | --- | --- |
| Calandri and colleagues , 2020 | Argentina | 40 (20 SA) | 36% female (45% female SA) | Cross-sectional observational study | SA were those who performed above -1 Standard Deviation (SD) of the mean on an episodic memory test (delayed recall memory test from the Rey Auditory Verbal Learning Test (RAVLT)) compared to those two decades younger (aged 50-60 years old) and who had a normal score for their age range on the Boston Naming Test (BNT); the Trail Making Test Part B (TMT-B), and the Semantic Fluency Test. |
| Cook Maher and colleagues , 2022 | USA | 79 (56 SA) | 72% female (73% female SA) | Longitudinal observational study (Northwestern University SuperAging Program) | SA were defined as adults over age 80 with delayed recall memory scores (RAVLT) above the mean for 56-64 year olds and at least average-for-age performance in non-episodic memory domains (BNT, TMT-B, and category fluency test). |
| Dang and colleagues , 2019 | Australia | 358 (179 SA) | 68% female (68% female SA) | Australian Imaging, Biomarkers and Lifestyle (AIBL) study | SA were those aged 60+ who performed above the sex-adjusted normative average for 30-44 year olds on a delayed recall memory test (California Verbal Learning Test – Second Edition (CVLT-II) Long Delay Free Recall trial), and above -1 SD using published normative data for all non-memory tests (assessing executive function, working memory, and processing speed). |
| Doyle and colleagues , 2024 | USA (only Latinx participants included) | 820 (76 SA) total, 640 (45 SA) from PREHCO and 180 (31 SA) | 64% female in PREHCO (71% female SA) and 69% female in HRS (71% female SA) | Assessed participants from two longitudinal observational studies the Puerto Rican Elder Health Conditions (PREHCO) study and Hispanic | Participants were considered SA if they were at least 80 years old at the most recent wave, scored at or above the median for immediate and delayed recall scores of Hispanic respondents aged 55-64 from the HRS, and had no cognitive impairment at all waves (assessed using the Mini Mental |
| García-Chanes and colleagues , 2022 | Mexico | 590 (74 SUMAs) | 55.6% female (66% female SUMAs) | Prospective cohort study (Mexican health and ageing study) | In this study SAs were referred to as successful memory agers (SUMAs) and were those aged over 75 years with memory performance within or above the normative values for subjects aged 65 to 69. Normative values of the “ 10-word learning test” from the CERAD protocol, included in the Mex-Cog battery, were used to assess memory. |

| Authors, year | Country | Sample Size<br>(No. SAs, No.<br>controls) | % female Total (%)<br>female SA) | Study Design (cohort if<br>applicable) | SuperAger Assessment |
| --- | --- | --- | --- | --- | --- |
| Garó-Pascual<br>and<br>colleagues,<br>2023 | Spain | 119 (64 SA) | 61% female (59%<br>female SA) | Prospective cohort study<br>(Vallecas project longitudinal<br>cohort) | SAs were those aged 80 years or older with the episodic memory scores (assessed using the free and Cued Selective Reminding Test) at or above the mean of those 20–30 years younger (50–56 years) at the same education level and who performed with $-1$ SD of their age group on non-memory tests (BNT-B and category fluency test). |
| Hermansen<br>and<br>colleagues,<br>2024 | Denmark | 1666 (27<br>CHPOOI) in<br>1905 cohort<br>and 1050 (33<br>CHPOOI) in<br>1915 cohort | 72.48% female (70.37%<br>female CHPOOI) in<br>1905 cohort and<br>72.95% female<br>(69.70% female<br>CHPOOI) in 1915<br>cohort | Danish 1905 and 1915 birth<br>cohorts | In this study SAs were referred to as Cognitively High-Performing Oldest Old Individuals (CHPOOI), being defined at those with cognitive test scores on the cognitive composite scores comprising of 5 tests (Fluency test, Digits Forward test, Digits Backward test, Immediate recall test, and Delayed Recall test) above the mean of middle-aged subjects (50–65 years old), and scores above 28 on the mini-mental state exam (MMSE). |
| Maccora and<br>colleagues,<br>2020 | Australia | 1679 (116 SA) | 49% females (61%<br>female SA) | Prospective cohort study<br>(Personality and Total Health<br>Through Life (PATH) Cohort<br>Study) | SAs were those from the 60s cohort who, for all three consecutive waves over a period of 12 years, maintained both immediate and delayed recall scores at or above the median of the immediate and delayed recall scores of participants of the same gender from the 20s cohort and an MMSE score of 29 or 30 for that wave. |
| Saint Martin<br>and<br>colleagues,<br>2017 | France | 543 (186 CE) | 59% female (53%<br>female CE) | Prospective cohort study (the<br>PROOF cohort) | In this study SAs were referred to as cognitively elite (CE) being those with scores above the mean for each of the three cognitive domains of information processing speed and attention (TMT- A, the Stroop Colour-Word Test (Parts I and II) and the Coding subtest of the Wechsler Adult Intelligence Scale, Third Edition (WAIS-III)), memory (the Benton Visual Retention Test (form C) and the Grober and Buschke Selective Reminding Test), and executive function (TMT- B, the Stroop Colour-Word Test (part III), an alphabet fluency test, and a category fluency test). |

| Authors, year | Country | Sample Size<br>(No. SAs, No.<br>controls) | % female Total (%<br>female SA) | Study Design (cohort if<br>applicable) | SuperAger Assessment |
| --- | --- | --- | --- | --- | --- |
| Trammell<br>and<br>colleagues,<br>2024 | USA (Only<br>African<br>Americans<br>recruited) | 1282 (61 SA) | 75% female (87%<br>female SA) | NIH- NIA-supported Alzheimer's<br>Disease Research Centers<br>(ADRCs) | SA were referred to as those with exceptional memory being those aged $\geq 80$ with normal cognition (MoCA scores) and episodic memory (craft story test of delayed recall) and non-memory (TMT-A & B) z-scores within one SD of the mean. |
| Wagner and<br>Grodstein,<br>2022 | USA | 2730 (455<br>exceptional<br>episodic<br>memory<br>agers) | 100% female | Prospective cohort study (NHS<br>cohort) | In this study SAs were referred to as those with exceptional episodic memory being those with scores $\geq 1.5$ SD above the mean on the immediate and delayed recall of the east Boston memory test and immediate and delayed recall on the verbal episodic memory test from the Telephone Interview of Cognitive Status (TICS). |
| Yang and<br>colleagues,<br>2022 | China | 1347 (154 SA) | 24% female (65%<br>female SA) | Prospective cohort study<br>(Beijing Aging Brain<br>Rejuvenation Initiative) | SA were those with excellent episodic memory or executive function, i.e., scoring higher than 1.5 SDs (SDs) above the age- and education-adjusted mean on the Auditory Verbal Learning Test long-time delayed recall (AVLT-N5) or the TMT-B; and intact general cognition, as evidenced the lack of a score on any test lower than 1.5 SDs below the age- and education-adjusted means. |
| Yu and<br>colleagues,<br>2020 | Singapore | 327 (64 SA) | 81% female SA (not<br>reported overall) | Prospective cohort study (Aging<br>in a Community Environment<br>Study cohort) | SA were those with superior cognitive functioning (i.e., index scores $> 115$) in at least one of the five Repeatable Battery for the Assessment of Neuropsychological Status (RBANS) domains; with at least average cognitive functioning (i.e., index scores $> 100$) in each of the other four RBANS domains; and intact daily functioning as defined by having a clinical dementia rating of 0. |

SA status was assessed using various criteria across studies, with a range of different synonymous terms such as “successful cognitive agers” and “cognitive elite” also referred to in several papers (Cook Maher et al., 2022; Garcia-Chanes et al., 2022). Several studies defined SAs using the original Northwestern University SuperAging programme definition of individuals aged 80 years or older with exceptional episodic memory performance, scoring higher than 1.5 SDs above the mean (Harrison et al., 2012). Others considered successful memory agers as those aged over 75 years with memory performance within or above normative values for younger age groups. Additional assessments included superior cognitive functioning, intact daily functioning, and maintaining cognitive scores above the mean for specific cognitive domains. Each study employed distinct criteria to identify SAs (or synonymous term, e.g., superior memory ager, cognitive elite), reflecting the diversity of approaches in defining this unique group of individuals, with some using less stringent criteria compared to the original SA definition, for instance comparing older adults to individuals aged 65-69 instead of middle aged adults (Garcia-Chanes et al., 2022; Garo-Pascual et al., 2023), or classifying SA as those who only performed above the mean on specific cognitive tests (Saint Martin et al., 2017). Details of study characteristics are presented in Table 1.2. Across all included studies, reported SAs tended to be proportionately more female than the overall study population, although this was only a statistically significant difference in two studies (Trammell et al., 2024a; Yang et al., 2022). In terms of education, eight of the included studies reported higher levels of education in the SAs sample compared with controls, although this was only a statistically significant difference in four of the studies (Doyle et al., 2024; Garcia- Chanes et al., 2022; Garo-Pascual et al., 2023; Hermansen et al., 2024; Maccora et al., 2021; Saint Martin et al., 2017; Trammell et al., 2024b; Yang et al., 2022), with Wagner and Grodstein (2022) and Dang and colleagues (2019) matching their subjects on education.

### 3.2 Risk of Bias in Studies

Two reviewers independently rated each study’s quality using the Quality of Prognosis Studies in Systematic Reviews (QUIPS) tool (Hayden et al., 2006). The QUIPS tool assesses six potential components of bias: inclusion, attrition, prognostic factor measurement, confounders, outcome measurement, and analysis and reporting. Disagreements in ratings were resolved by consensus, with a third reviewer available to resolve any lack of agreement. The risk of bias of all the included studies is presented in Table 1 in the supplementary materials.

### 3.3 Results of individual studies/Lifestyle Factor Results

#### 3.3.1 Physical Activity

Seven studies explored the association between physical activity and SA status (see Table 1.3). The sample sizes of these studies varied widely, ranging from 40 to 2730 participants. Physical activity was assessed using diverse measures across the studies. Some studies utilized measures of sedentarism in daily life (Calandri et al., 2020), while others employed summed measures based on self-reported engagement in physical activities and the duration or frequency of each activity (Wagner & Grodstein, 2022). Most studies did not find significant differences in reported physical activity engagement between SAs and controls. However, the largest study (Wagner & Grodstein, 2022), observed consistently higher levels of physical activity among females with exceptional episodic memory from mid-life (aged 52-62) through to late life (80+ years). However, these levels did not differ significantly between mid-life and late life.

**Table 1.3.** Qualitative summary – Physical Activity in SAs.

| Author, year | N (N SAs) | Physical Activity measure(s) | Statistical Test | Outcome |
| --- | --- | --- | --- | --- |
| Calandri and colleagues, 2020 | 40 (20 SA) | Physical activity was measured according to the WHO criteria for sedentarism in daily life (sedentary or not) | *t*-test | No significant difference between SA and healthy controls |
| Garó-Pascual and colleagues, 2023 | 119 (64 SA) | Self-report weekly exercise frequency and duration | Random forest analysis (total 89 predictors considered) | No significant difference between SA and healthy controls on either exercise measure |
| Maccora and colleagues, 2020 | 1679 (116 SA) | Exercise recorded as % participation in none/mild, moderate and vigorous exercise per week (as per Whitehall II study) | *t*-test | No significant difference between SA and healthy controls in either men or women |
| Saint Martin and colleagues, 2017 | 543 (186 CE) | Physical Activity Index score [0-9] | Mann-Whitney or chi-squared, or ANOVA | No significant difference between CE and healthy controls |
| Wagner and Grodstein, 2022 | 2730 (455 exceptional episodic memory agers) | Through questionnaire the sum of all reported activities denoted total physical activity and the sum of walking and | For each lifestyle factor, mean trajectories were estimated using | Higher levels of PA associated with being an exceptional episodic memory ager. |
|  |  | stair-climbing denoted low-intensity physical activity, expressed in metabolic equivalents (MET)-h/week | latent process mixed models |  |
| Yang and colleagues, 2022 | 1347 (154 SA) | PA assessed as part of leisure activity questionnaire, weight grade method used to calculate PA score | ANOVA, multivariable regression analysis, and structural equation model mention considering early-life cognitive reserve (education and occupational attainment) and late-life leisure activities (mental, physical, and social activities) | No significant difference between SA and healthy controls only between SA and cognitively impaired |
| Yu and colleagues, 2020 | 327 (64 SA) | Questionnaire assessing PA frequency (e.g., Never or rarely, more than once a month but less than once a week, one to three times a week, four to six times a week, and daily) | A latent class analysis | PA did not show an association with SA status |

Overall, the findings regarding the relationship between physical activity and SuperAger status are mixed, with further inconsistencies in the measurement of physical activity. Further research is warranted to better understand the relationship between physical activity and SuperAger status, with particular emphasis on elucidating any potential causal mechanisms, such as whether increased or regular physical activity contributes to enhanced cognitive functioning that characterizes SuperAgers. This highlights the need for the development and utilization of a standardized and comprehensive scale to accurately measure physical activity levels in future investigations.

#### 3.3.2 Social Engagement

Seven studies reported data on social activity engagement (see Table 1.4); a variety of measures were used as indicators of social activity engagement. Several studies assessed the number of social activities in a week (García-Chanes et al., 2022; Maccora et al., 2021), as well as formal questionnaires including the social activity index (Saint Martin et al., 2017), and the leisure activity scale (Yang et al., 2022).

**Table 1.4.** Qualitative summary – Social engagement in SAs.

| Authors, year | N (N SAs) | Social Engagement measure | Statistical test | Outcome |
| --- | --- | --- | --- | --- |
| Calandri and colleagues, 2020 | 40 (20 SA) | Measured through the Social Network Index (Network size and Diversity sub-scores) | *t*-test or Mann-Whitney | No significant difference between SA and controls |
| García-Chanes and colleagues, 2022 | 590 (74 SA) | Binary variable created for participation in 2 or more activities (out of list of 9) | Chi-squared test and multinomial logistic regressions | A significant *p*-value was observed between SA status and participating in 2+ social activities, although social engagement was not a significant predictor of SA status in the multinomial logistic regression |
| Garo-Pascual and colleagues, 2023 | 119 (64 SUMAs) | Questionnaire that assessed several social engagement measures including composite sociality, composite mental use, and composite midlife activities | Random forest analysis (total 89 predictors considered) | Only midlife activities and musical background were associated with significantly increased SUMA odds |
| Maccora and colleagues, 2020 | 1679 (116 SA) | Questionnaire assessing number of social activities in a week and number of investigative activities in a week | *t*-test and logistic regression | For women, more participation in investigative activities associated with statistically significant increased SA odds. For men, more social activities were also significant SA predictors |
| Saint Martin and colleagues, 2017 | 543 (186 CEs) | Social Activity Index questionnaire | Kruskal-Wallis or ANOVA and multiple logistic regression | Statistically significantly higher levels of social activity were observed in CEs. |
| Yang and colleagues, 2022 | 1347 (154 SA) | Scale of 23 items recognised as common leisure activities in the older Chinese population. Weighted grade method used to calculate scores for mental, physical, and social activities individually | ANOVA, multivariable regression analysis, and structural equation model considering early-life cognitive reserve (education and occupational attainment) and late-life leisure activities (mental, physical, and social activities) | A significant difference was observed between SA and cognitively impaired individuals but not between cognitively normal individuals and SA |
| Yu and colleagues, 2020 | 327 (64 SA) | Questionnaire assessing weekly frequency of social activity engagement | Kruskal-Wallis and Latent class analysis | Social isolation (as opposed to social engagement) was statistically associated with superior cognition |

The results for social engagement were mixed across these studies. Two studies reported higher levels of social engagement in SAs compared to healthy controls (García-Chanes et al., 2022; Saint Martin et al., 2017), whereas one study found significant differences only between SAs and cognitively impaired individuals (Yang et al., 2022). Garo-Pascual and colleagues (2023) observed a significant effect only in midlife social activities, not in later life. Maccora and colleagues (2020) reported gender-specific findings, with female SAs showing higher engagement in investigative activities and male SAs exhibiting greater social activity engagement compared to controls.

Calandri and colleagues (2020) did not find a relationship between SA status and social network size. It is important to note that this measure differs substantially from other indicators of social engagement, and their study had a small sample size of 40 participants. Conversely, Yu and colleagues (2020) found a significant association between social isolation and SA status in later life, although this finding was not consistently supported by other studies.

#### 3.3.2 Diet

Two studies reported data on the relationship between diet and SA status (see Table 1.5). Both studies utilized Mediterranean diet scores to measure dietary quality, with Wagner and Grodstein (2022) further examining dietary patterns from mid-life to later life. Neither study found evidence supporting a significant relationship between diet and SA status in older adults. Garo-Pascual and colleagues (2023) also investigated the association between dietary habits in midlife and SA status in older age, finding no significant correlation.

**Table 1.5.** Qualitative summary – Diet in SAs.

| Authors, year | N (N SAs) | Diet measure(s) | Statistical test | Outcome |
| --- | --- | --- | --- | --- |
| Garó-Pascual and colleagues, 2023 | 119 (64 SA) | 13 different dietary pattern measures (e.g., dairy consumption) including Mediterranean diet score | Random forest analysis (total 89 predictors considered) | No significant difference between SA and controls |
| Wagner and Grodstein, 2022 | 2730 (455 exceptional episodic memory agers) | Mediterranean diet score assessed from mid to later life | For each lifestyle factor, mean trajectories were estimated using latent process mixed models | No significant difference between exceptional episodic memory agers and controls in mid or later life |

#### 3.3.3 Alcohol

Six studies looked at the association between SA status and alcohol consumption (see Table 1.6). One study reported that higher alcohol consumption among women was linked to increased odds of SA status, although this association was not evident among men (Maccora et al., 2021). While another study conversely observed lower rates of alcohol consumption in a sample of African American SAs (Trammell et al., 2024a). One more study reported that CHPOOI individuals were more likely to be drinkers in one of the two cohorts they examined (Hermansen et al., 2024). Moreover, three studies found no significant association between alcohol consumption and SA status (Garo- Pascual et al., 2023; Saint Martin et al., 2017; Yu et al., 2020).

**Table 1.6.**
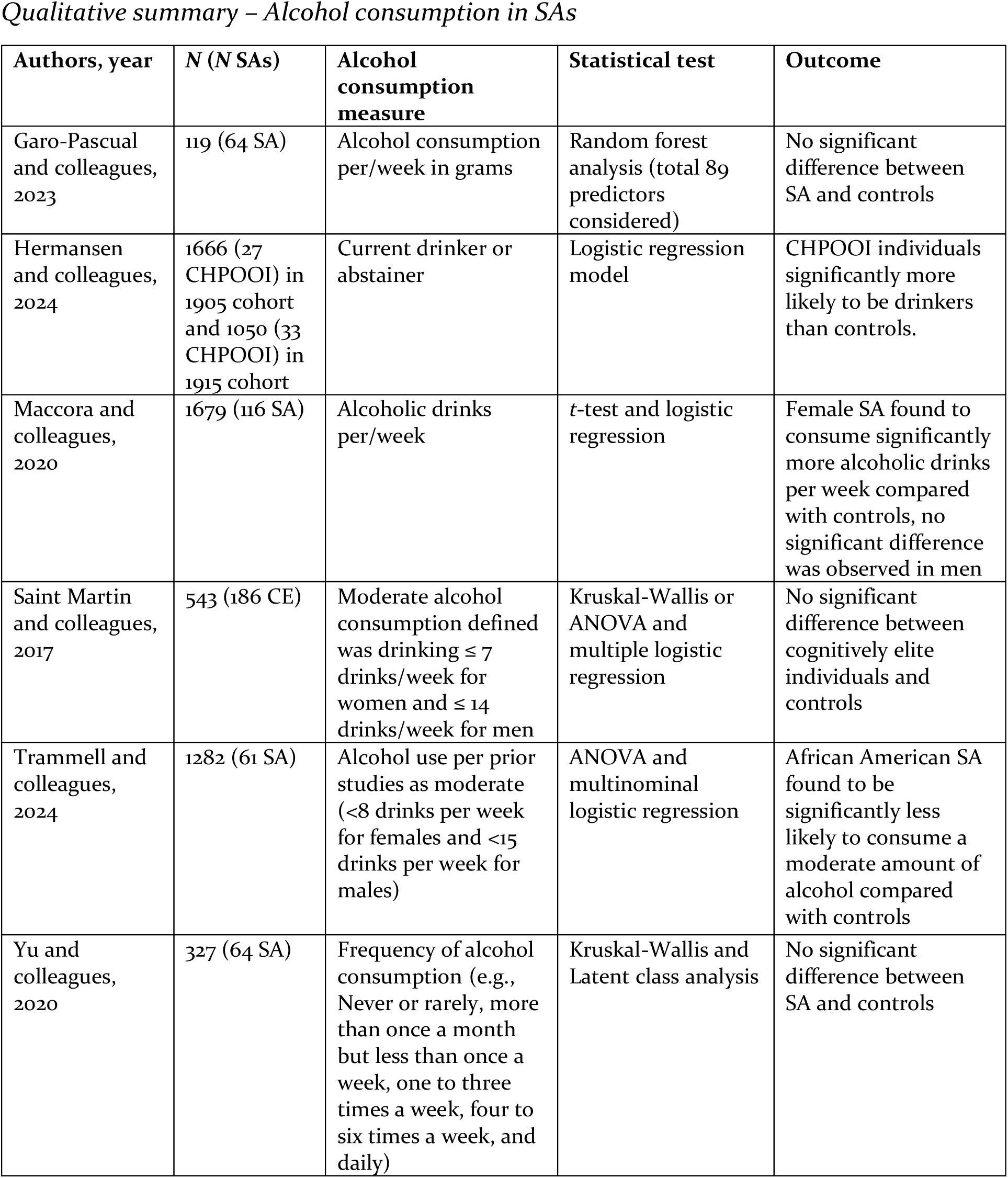
Qualitative summary – Alcohol consumption in SAs.

#### 3.3.4 Smoking

Eight studies examined the association between SA status and smoking (see Table 1.7). Smoking was measured by smoking history in three studies and by current smoker status in five studies. Across all studies examining current smoking status, no significant associations were observed between SAs and controls. Among the studies investigating smoking history, only one reported a significant finding: Saint Martin and colleagues (2017) observing a significant difference between CE individuals who maintained their cognition at follow-up 7-10 years later (stable CE) and CE individuals who showed worsened cognition, however, no difference was observed between the stable CE group and stable cognitively normal group

**Table 1.7.**
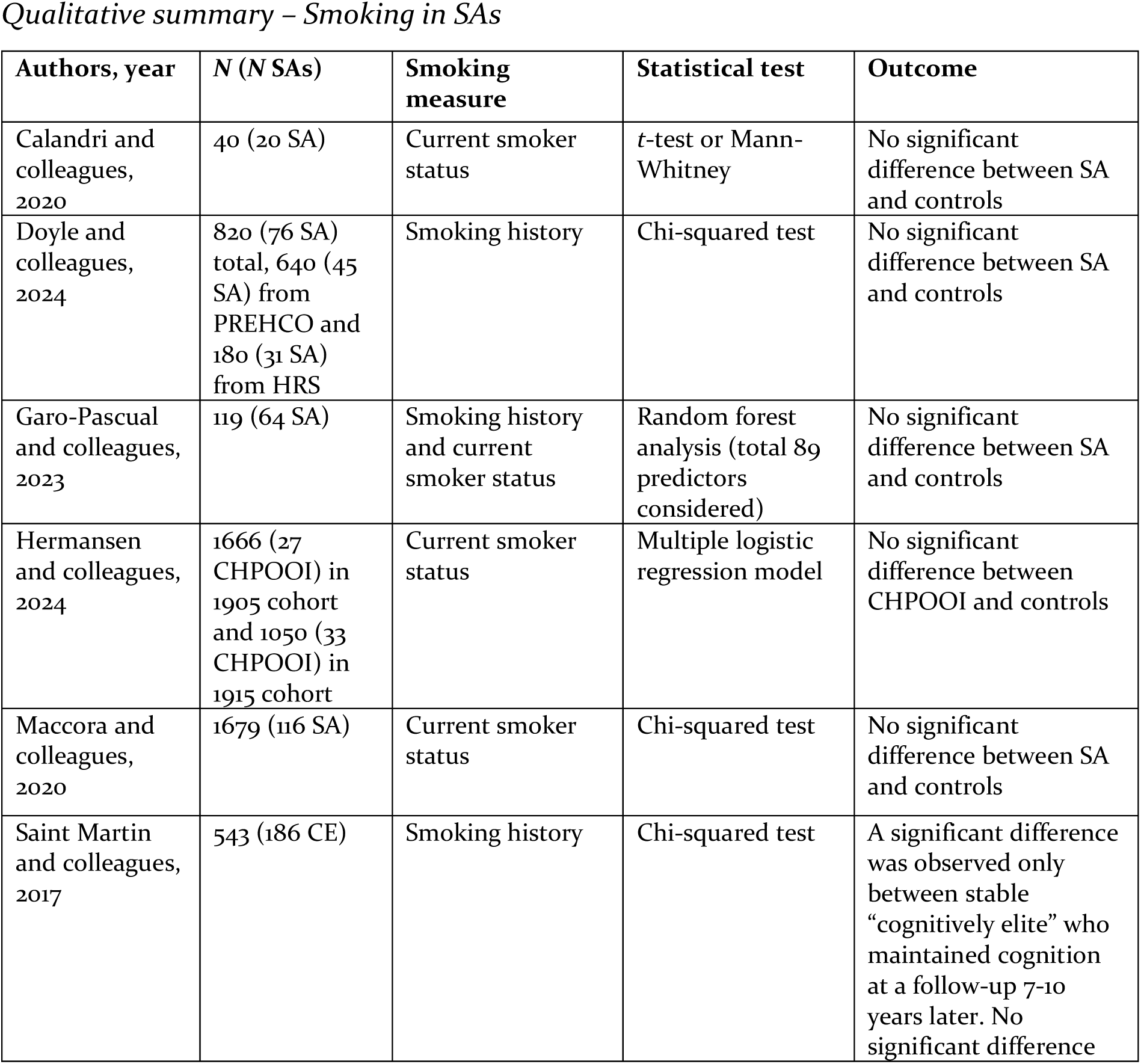
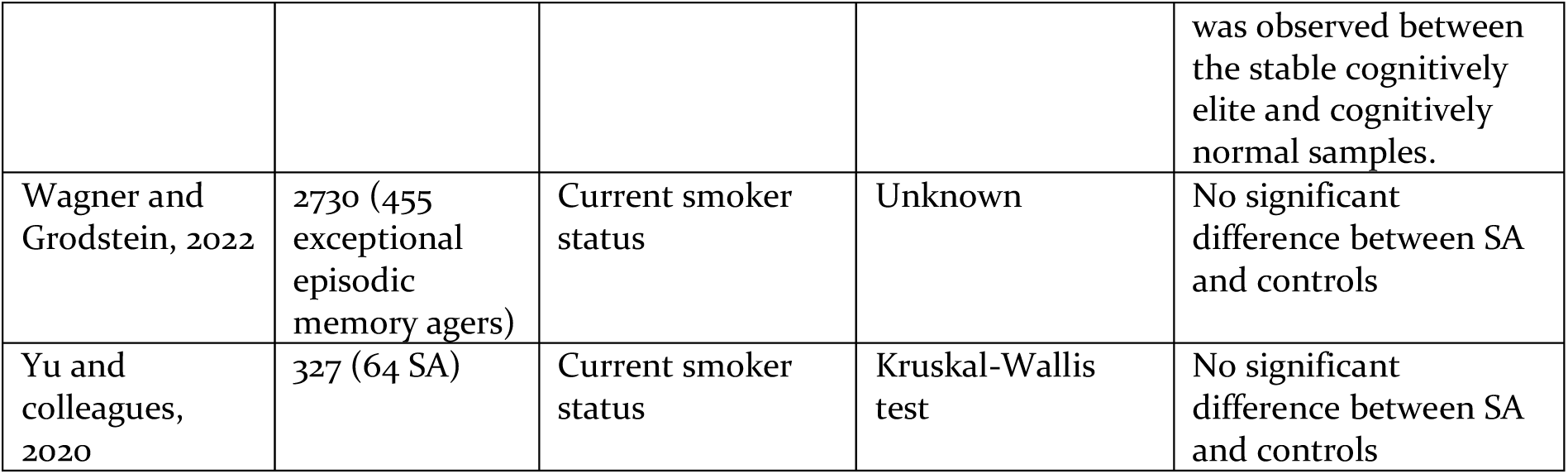
Qualitative summary – Smoking in SAs.

| **Authors, year** | **N (N SAs)** | **Smoking measure** | **Statistical test** | **Outcome** |
| --- | --- | --- | --- | --- |
| Calandri and colleagues, 2020 | 40 (20 SA) | Current smoker status | *t*-test or Mann-Whitney | No significant difference between SA and controls |
| Doyle and colleagues, 2024 | 820 (76 SA) total, 640 (45 SA) from PREHCO and 180 (31 SA) from HRS | Smoking history | Chi-squared test | No significant difference between SA and controls |
| Garo-Pascual and colleagues, 2023 | 119 (64 SA) | Smoking history and current smoker status | Random forest analysis (total 89 predictors considered) | No significant difference between SA and controls |
| Hermansen and colleagues, 2024 | 1666 (27 CHPOOI) in 1905 cohort and 1050 (33 CHPOOI) in 1915 cohort | Current smoker status | Multiple logistic regression model | No significant difference between CHPOOI and controls |
| Maccora and colleagues, 2020 | 1679 (116 SA) | Current smoker status | Chi-squared test | No significant difference between SA and controls |
| Saint Martin and colleagues, 2017 | 543 (186 CE) | Smoking history | Chi-squared test | A significant difference was observed only between stable “cognitively elite” who maintained cognition at a follow-up 7-10 years later. No significant difference was observed between the stable cognitively elite and cognitively normal samples. |
| Wagner and Grodstein, 2022 | 2730 (455 exceptional episodic memory agers) | Current smoker status | Unknown | No significant difference between SA and controls |
| Yu and colleagues, 2020 | 327 (64 SA) | Current smoker status | Kruskal-Wallis test | No significant difference between SA and controls |

#### 3.3.5 Mental Health

Eleven studies reported data on the relationship between mental health and SA status (see Table 1.8). Mental health was measured using several metrics across these studies, 10 studies reported data on depression, five studies reported on anxiety and one study reported on stress. Most frequently, researchers utilized standardized scales, with the Geriatric Depression Scale (GDS; Yesavage et al., 1982), being the predominant choice, to measure mental health (see Table 1.8). Alternatively, some studies investigated participants’ self-reported history of depression or anxiety (Garo-Pascual et al., 2023; Saint Martin et al., 2017; Trammell et al., 2024a).

**Table 1.8.** Qualitative summary – Mental health in Sas.

| **Authors, year** | **N (N SAs)** | **Mental Health Measure** | **Statistical test** | **Main findings** |
| --- | --- | --- | --- | --- |
| Cook Maher and colleagues, 2022 | 79 (56 SA) | Short-form Geriatric Depression Scale | Analysis of covariance (ANCOVA) | No significant difference between SA and controls |
| Dang and colleagues, 2024 | 358 (179 SA) | Hospital Anxiety and Depression Scale | One way ANOVA | No significant difference between SA and controls |
| Doyle and colleagues, 2024 | 820 (76 SA) total, 640 (45 SA) from PREHCO and 180 (31 SA) from HRS | Center for epidemiologic studies depression (CES-D) scale, half of sample also received the “leave behind” questionnaire with anxiety assessment | *t*-test and multivariate logistic regression | SA showed significantly lower depressive symptoms compared with controls in both cohorts. SA in the PREHCO cohort also showed significantly lower anxiety scores compared to controls from PREHCO. |
| García-Chanes and colleagues, 2022 | 590 (74 SUMAs) | Self-report 2 questions on depression | Chi-squared test and multinomial logistic regressions | SA had significantly lower scores on the two depression questions compared with controls |
| Garo-Pascual and colleagues, 2023 | 119 (64 SA) | Geriatric depression scale, depression/anxiety history, STAI state and trait | Random forest analysis (total 89 predictors considered) | SA had significantly lower GDS scores and STAI state and trait scores, although no difference in anxiety and depression history rates compared with controls |
| Hermansen and colleagues, 2024 | 1666 (27 CHPOOI) in 1905 cohort and 1050 (33 CHPOOI) in 1915 cohort | Cambridge Mental Disorders of the Elderly Examination (CAMDEX) used to assess depression | Linear regression models | CHPOOI individuals had significantly lower depression scores than controls |
| Maccora and colleagues, 2020 | 1679 (116 SA) | Goldberg depression scale/Goldberg anxiety scale (GAS) | *t*-test and logistic regression | SA had significantly lower Goldberg Depression Scale scores in men but not women. No difference was seen on the GAS. |
| Saint Martin and colleagues, 2017 | 543 (186 CE) | History of anxiety/depression | Chi-squared test | No significant difference between “cognitively elite” and controls |
| Trammell and colleagues, 2024 | 1282 (61 SA) | History of depression/antidepressant use | ANOVA and multinomial logistic regression | Significantly lower depression history levels observed in African American SA compared with controls |
| Yang and colleagues, 2022 | 1347 (154 SA) | Geriatric depression scale | ANOVA | No significant difference between SA and controls |
| Yu and colleagues, 2020 | 327 (64 SA) | Frequency of feeling stressed | Kruskal-Wallis test | SA reported significantly higher stress levels than controls |

Five of the 10 studies investigating depression reported significant findings (see Table 1.8). In one study, the effect was observed in men and not women (Maccora et al., 2021), while another study measured depression using two self-report questions (Garcia- Chanes et al., 2022). Only one of the studies that examined history of depression reported any significant differences (Trammell et al., 2024).

Of the studies investigating anxiety, one study reported significantly lower anxiety scores in SAs on the “leave behind” questionnaire (Doyle et al., 2024), while another study observed a significant relationship between lower scores on the state trait anxiety inventory and SA status (Garo-Pascual et al., 2023). However, no such effect was observed between anxiety history and SA status in the same study, while three other studies reported no differences (Dang et al., 2019; Maccora et al., 2021; Saint Martin et al., 2017). In Yu and colleagues (2020) study reporting on stress, they observed that SAs reported feeling stressed significantly more often than controls.

#### 3.3.6 Sleep

Five studies provided data on sleep (see Table 1.9). Sleep was assessed across studies through various methods, including specific evaluations of sleep duration (Yu et al., 2020), sleep problems (Maccora et al., 2020), disorders (Trammell et al., 2024) and using specific questionnaires like the Pittsburgh Sleep Quality Index (Yang et al., 2022).

**Table 1.9.**
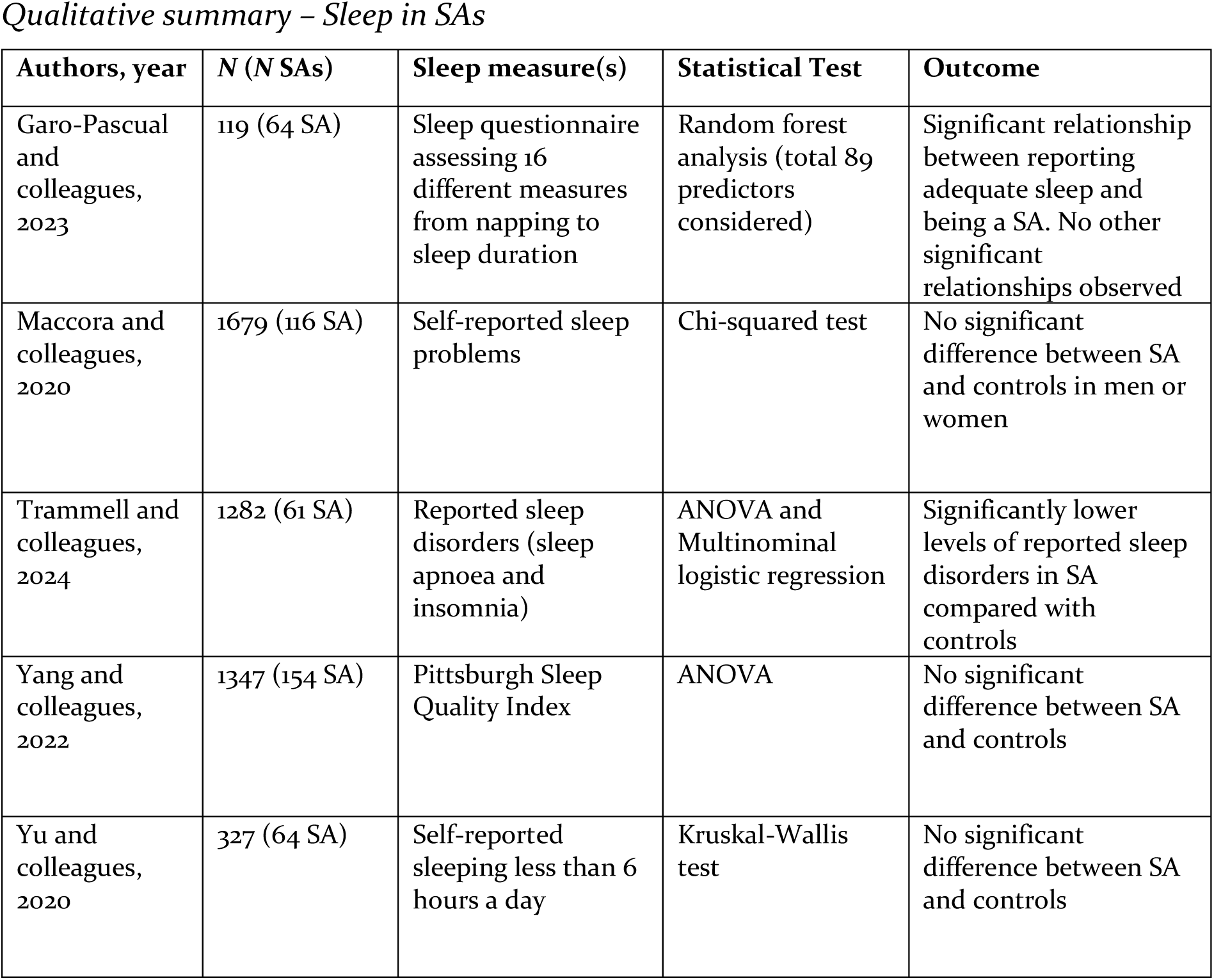
Qualitative summary – Sleep in SAs.

Consistently, three studies found no significant relationship between measures of sleep quality and SA status (Maccora et al., 2021; Yang et al., 2022; Yu et al., 2020). Conversely, Garo-Pascual and colleagues (2023) found a significant association between SA status and adequate sleep, while the other sleep measures did not yield significant results and Trammell and colleagues (2024) in a sample of older African American participants observed significantly lower levels of reported sleep disorders in SAs compared with cognitively normal controls.

## 4. Discussion

This review summarised the existing literature on SA, examining the potential effects that modifiable lifestyle factors may play to the cognitive preservation of SA. This review included 13 studies with seven different lifestyle factors considered (physical activity, social engagement, diet, alcohol, smoking, mental health, & sleep). A complicated picture was observed across these studies dependent on the lifestyle factor examined.

### 4.1 Physical Activity and SuperAger Status

Multiple systematic reviews and meta-analysis examining healthy and impaired cognitive ageing have suggested that regular exercise can protect against age-related cognitive decline (Blondell et al., 2014; Iso-Markku et al., 2024; Sofi et al., 2011). In the present study we observed very mixed results, with no clear associations between physical activity and the preservation of cognition in SA. While most studies did not report an association between physical activity and SA status, the inconsistency in the measurement of physical activity across these studies and the wide differences in sample size make any meaningful conclusions difficult to draw. Thus, the lack of significant results in the current study may more reflect the small sample size, study numbers, and heterogeneity in physical activity assessment. This review further highlights the need to establish standardised and comprehensive scale to accurately measure physical activity levels in future studies, with more studies needed to clarify and potential relationship between physical activity and SA status.

Further, previous research has tended to show that the beneficial effects of exercise may be modified by the age and gender of the sample, such that men may experience greater induced cognitive benefits earlier in life, while women exhibit greater benefits later in the lifespan. This finding could also explain Wagner and Grodstein’s (2022) observation that women with exceptional episodic memory had significantly higher physical activity levels. However, Maccora and colleagues (2020) did not observe a significant effect of late-life physical activity in male and female SAs challenging this theory and warranting further longitudinal investigations to examine how the timing of lifestyle interventions affects cognitive outcomes across the lifespan in men and women.

### 4.2 Social Engagement and SuperAger Status

Most studies included in our review suggest that social engagement may positively contribute to SA status. This aligns with previous research on the relationship between social engagement and dementia risk, with two systematic reviews reporting that less frequent social contact was associated with a higher risk of dementia (Kuiper et al., 2015; Penninkilampi et al., 2018). Similarly, a more recent systematic review observed a positive effect of leisure and social activities in seven of the eight studies examined (Ye et al., 2023).

However, some studies did not find significant results, while others reported only partially significant findings. These divergent outcomes may be due to the variety of measures used to assess social engagement, making it challenging to draw definitive conclusions about the relationship between SA status and social engagement.

Notably, the studies that reported significant associations between SuperAger status and social engagement all used measures that specifically assessed social activity engagement. This suggests that social activity engagement may play a role in contributing to SuperAger status. However, this trend is not universally supported, as Yu and colleagues (2020) found that social isolation was associated with SuperAger status. While this finding may be a statistical anomaly, it warrants further investigation to better understand the nuances of the relationship between social engagement and SuperAger status and the way SA status was assessed.

### 4.3 Diet and SuperAger Status

Previous research on diet has shown that it may be an important factor in the prevention of dementia and the improvement of cognition. In particular, the Mediterranean diet has garnered considerable attention in studies of modifiable lifestyle factors and cognition, with previous research demonstrating its association with beneficial effects on various health-related outcomes, including cognitive function (Bhushan et al., 2018; Maggi et al., 2023; Nucci et al., 2024; Shannon et al., 2019; Tangney et al., 2014). However, while several studies have reported positive effects of adherence to the Mediterranean diet, not all research has supported these findings (Charisis et al., 2021; Glans et al., 2023; Haring et al., 2016; Kesse-Guyot et al., 2013). Some studies have found no protective effects, and the Lancet 2024 Commission on Dementia concluded that there is insufficient evidence to consider it a definitive risk factor for dementia (Livingston et al., 2024).

In the two studies included in this review examining the relationship between adherence to the mediterranean diet and SA status, both failed to find a significant relationship (Garo-Pascual et al., 2023; Wagner & Grodstein, 2022). This might suggest that perhaps diet may not be an important contributory factor to the preservation of cognitive abilities in SAs. However, there are other potential reasons for these results that should be considered before making any firm conclusions on this relationship. Firstly, this review included only two studies that investigated diet and SA status. Additionally, both studies focused exclusively on the Mediterranean diet, which, while prominent in epidemiological literature and widely studied in relation to cognition, is not the only diet with potential cognitive benefits.

### 4.4 Alcohol and SuperAger Status

High alcohol consumption has been shown across multiple studies to increase the risk of several chronic diseases including cognitive impairment and dementia (Deng et al., 2006; Hulse et al., 2005; Jeon et al., 2023; Langballe et al., 2015; Zarezadeh et al., 2024), with high alcohol consumption considered a risk factor for dementia by the 2024 Lancet Commission on dementia risk (Livingston et al., 2024).

However, in three of the six studies included in this review, alcohol consumption was not significantly associated with SA status (Garo-Pascual et al., 2023; Saint Martin et al., 2017; Yu et al., 2020). While three studies did find significant relationships between alcohol consumption and SA status, these results were in the opposite direction and should be interpreted with caution. For instance, Maccora and colleagues (2021) reported that female SAs consumed significantly more alcoholic drinks per week compared to controls, but they advised caution due to methodological challenges, such as the reliability of self-reported alcohol consumption and the influence of confounding variables like socioeconomic status and overall health (Anstey & Peters, 2018; Maccora et al., 2021). Further, Hermansen and colleagues (2024) reported that CHPOOI individuals were significantly more likely to drink in one of the two cohorts they examined, however, considering this study only used a crude measure of alcohol consumption (drinker vs abstainers) and that it was only observed in one cohort it should be treated with caution.

In one further study in our review examining African American SAs, they observed lower levels of alcohol consumption among SA, although these findings could be a reflection on cultural differences with further research needed examining SAs in different ethnic and cultural groups. In summary, while this body of evidence suggests that alcohol consumption may not play a significant role in the preservation of cognitive abilities in SAs, further research is needed before drawing firm conclusions.

### 4.5 Smoking and SuperAger Status

Smoking is another factor which was considered a risk factor for dementia by the 2024 Lancet Commission on dementia risk (Livingston et al., 2024). However, smoking was not found to be a significant predictor of SA status in the studies we reviewed. With both studies examining current smoker status and smoking history failing to observe a significant interaction. Thus, it may be the case that while a risk factor for dementia, smoking is less important for SAs preserved healthy cognitive abilities.

### 4.6 Mental Health and SuperAger Status

Poor mental health has been identified as a risk factor for dementia across multiple studies, with a number of studies and reviews finding a significant link between mental disorders such as depression and anxiety with dementia and MCI (Gulpers et al., 2016; Ismail et al., 2017; Stafford et al., 2022). However, the present review found mixed results in studies examining depression, anxiety, and stress. One possible explanation for these inconsistent findings is the methodological variability among the included studies, particularly in the assessment tools used to measure mental health, as well as differences in sample characteristics and study aims.

For instance, most studies investigating the history of depression and/or anxiety, except for one (Trammell et al., 2024), reported no statistically significant differences between SAs and healthy controls on these measures (Garo-Pascual et al., 2023; Saint Martin et al., 2017). However, in studies that used formal depression scales, such as the GDS or CES-D, SAs were generally found to have significantly lower scores compared to cognitively healthy controls (Doyle et al., 2024; Garo-Pascual et al., 2023; Maccora et al., 2021). This suggests that current mental health status, as reflected by lower depression scores, may play a more important role in the preservation of cognitive function in SAs than a history of mental health conditions.

In studies examining anxiety, the findings were more mixed. Half of the included studies reported significantly lower anxiety scores in SAs (Doyle et al., 2024; Garo-Pascual et al., 2023), while the other half found no statistical difference between SAs and controls (Dang et al., 2019; Maccora et al., 2021; Saint Martin et al., 2017). This inconsistency could be attributable to the diversity of anxiety assessment tools used, as each study employed different questionnaires. Without further research using standardized measures of anxiety, it is difficult to draw definitive conclusions about the relationship between anxiety and SA status.

Finally, one study that examined stress found that SAs reported higher levels of stress compared to controls (Yu et al., 2020). This was the only study to investigate stress and the only one to find a negative correlation between mental health outcomes and SA status. Given that this is an isolated finding, more research is needed to explore the potential relationship between stress and SA status.

### 4.7 Sleep and SuperAger Status

Previous research examining the potential role of sleep in cognitive decline and dementia has yielded mixed results. The most recent 2024 Lancet Commission on Dementia Prevention concluded that there is insufficient consistent evidence to establish sleep as a modifiable risk factor for dementia (Livingston et al., 2024). Specifically, the commission noted that it remains unclear whether too little sleep (typically defined as less than 5 hours) or too much sleep (more than 10 hours) is associated with an increased risk of dementia or if these patterns reflect the prodromal stage of the disease (Livingston et al., 2024).

In the present review, a similarly mixed picture emerged across the included studies. Three studies reported no relationship between sleep and SA status, while two studies found significant associations. Of the two studies reporting significant findings (Garo-Pascual et al., 2023; Trammell et al., 2024a), one observed significantly lower levels of sleep disorders among SAs compared to controls (Trammell et al., 2024a). The other study reported that SAs were significantly more likely to report having adequate sleep, although no significant differences were found in other sleep metrics such as sleep duration or napping (Garo-Pascual et al., 2023).

Among the studies that found no difference between sleep and SA status (Maccora et al., 2021; Yang et al., 2022; Yu et al., 2020), one observed no difference in the proportion of SA and controls reporting insufficient sleep (less than 6 hours per night, Yu et al., 2020), another found no significant differences in Pittsburgh Sleep Quality Index scores between the two groups (Yang et al., 2022), and a third found no difference in self-reported sleep problems (Maccora et al., 2021).

A major issue in the literature on sleep and cognitive outcomes, which is also evident in the studies included in this review, is the diversity of measures used to assess sleep. The five studies in this review employed various methods, including measures of sleep duration, sleep disorders, and sleep quality, contributing to the inconsistencies in the findings. Therefore, the current disparate results likely reflect the heterogeneity of sleep measures used across studies.

In conclusion, given the heterogeneity and limited number of studies exploring the relationship between sleep and SA status, it is too early to draw definitive conclusions. Further research is needed, specifically studies that examine sleep duration, sleep disorders, and sleep quality, to better understand the potential relationships between sleep and SA status.

### 4.8 Limitations and Strengths

The present review is not without limitations. First, the studies included employed various cognitive function tools to define SAs, and different definitions of SA status—or synonymous terms—were used across studies. Second, the studies applied varying definitions to individual lifestyle factors and their combinations, which may have contributed to the heterogeneity of the findings. The development of standardized measures for different lifestyle factors could help address this issue. Third, most of the studies included were cross-sectional, which limited the ability to examine how lifestyle factors change across the lifespan or to establish causality in the relationship between these factors and SA status. Lastly, there was a limited number of studies investigating modifiable lifestyle factors in SAs. This review underscores the need for further research examining the influence of multiple lifestyle factors on SA status to better understand their role.

Despite these limitations, this review also has notable strengths. First, it is the first known review to summarize the association between multiple lifestyle factors and cognitive function in SAs. Second, the review included studies from a wide range of countries, with participants from diverse ethnic backgrounds, enhancing the generalizability of the findings. Third, the present review also took into consideration and evaluated how SA was defined in the included studies of this review.

### 4.9 Conclusions

In conclusion, this review highlights the potential influence of modifiable lifestyle factors—such as physical activity, social engagement, diet, alcohol consumption, smoking, mental health, and sleep—on SA status. While social engagement showed more consistent associations, evidence for other factors remains inconclusive due to methodological variability. More standardized, longitudinal studies are needed to confirm these relationships and to clarify the role of lifestyle in cognitive preservation among SAs. Promoting a healthy lifestyle may be key to supporting cognitive resilience, though further research is essential to establish stronger, evidence- based guidelines.

## Data Availability

N/A

