## Supplemental Table 1 for "Determining the role of lifestyle factors in healthy cognitive preservation in SuperAgers: A systematic review"

### Supplementary Materials

Table 1

Risk of bias of included studies of this review assessed using the QUIPS tool

| Author | Study participation | | | | Study attrition | | | | Prognostic factor measurement | | | | Outcome measurement | | | | Confounding measurement and account | | | | Analysis | | | | Decision |
|---|---|---|---|---|---|---|---|---|---|---|---|---|---|---|---|---|---|---|---|---|---|---|---|---|---|
| | Yes | Partly | No | Unsure | Yes | Partly | No | Unsure | Yes | Partly | No | Unsure | Yes | Partly | No | Unsure | Yes | Partly | No | Unsure | Yes | Partly | No | Unsure | |
| Calandri and colleagues, 2020 | | √ | | | √ | | | | | √ | | | | √ | | | | √ | | | | √ | | | Include |
| Cook Maher and colleagues, 2022 | √ | | | | √ | | | | √ | | | | √ | | | | √ | | | | √ | | | | Include |
| Dang and colleagues, 2019 | | √ | | | √ | | | | √ | | | | | √ | | | | √ | | | | √ | | | Include |
| Doyle and colleagues, 2024 | √ | | | | | √ | | | | √ | | | | √ | | | √ | | | | | √ | | | Include |
| García-Chanes and colleagues, 2022 | √ | | | | √ | | | | | √ | | | | √ | | | √ | | | | | √ | | | Include |
| Garo-Pascual and colleagues,2023 | √ | | | | | √ | | | | √ | | | √ | | | | √ | | | | | √ | | | Include |
| Hermansen and colleagues, 2024 | √ | | | | √ | | | | | √ | | | | √ | | | √ | | | | | √ | | | Include |
| Maccora and colleagues, 2020 | √ | | | | √ | | | | √ | | | | √ | | | | | √ | | | √ | | | | Include |
| Saint Martin and colleagues, 2017 | √ | | | | √ | | | | √ | | | | √ | | | | | √ | | | | √ | | | Include |
| Trammell and colleagues, 2024 | √ | | | | | √ | | | √ | | | | | √ | | | | √ | | | | √ | | | Include |
| Wagner and Grodstein, 2022 | | √ | | | √ | | | | √ | | | | √ | | | | √ | | | | √ | | | | Include |
| Yang and colleagues, 2022 | √ | | | | √ | | | | | √ | | | | √ | | | | √ | | | √ | | | | Include |
| Yu and colleagues, 2019 | √ | | | | | √ | | | √ | | | | | √ | | | | √ | | | √ | | | | Include |
